# Direct comparison of clinical diagnostic sensitivity of saliva from buccal swabs versus combined oro-/nasopharyngeal swabs in the detection of SARS-CoV-2 B.1.1.529 Omicron

**DOI:** 10.1101/2023.01.05.23284222

**Authors:** Andreas Puyskens, Janine Michel, Anna Stoliaroff-Pépin, Fatimanur Bayram, Akin Sesver, Ole Wichmann, Thomas Harder, Lars Schaade, Andreas Nitsche, Caroline Peine

**Affiliations:** Highly Pathogenic Viruses, Centre for Biological Threats and Special Pathogens, WHO Reference Laboratory for SARS-CoV-2 and WHO Collaborating Centre for Emerging Infections and Biological Threats, Robert Koch Institute, Berlin, Germany; Department for Infectious Disease Epidemiology, Immunization Unit, Robert Koch Institute, Berlin, Germany

**Keywords:** SARS-CoV-2, B.1.1.529 (Omicron), clinical diagnostic sensitivity, saliva, buccal swab sampling

## Abstract

While current guidelines recommend the use of respiratory tract specimens for the direct detection of SARS-CoV-2 infection, saliva has recently been suggested as preferred sample type for the sensitive detection of SARS-CoV-2 B.1.1.529 (Omicron). Here, we compare the clinical diagnostic sensitivity of paired buccal saliva swabs and combined oro-/nasopharyngeal swabs from hospitalized, symptomatic COVID-19 patients collected at median six days after symptom onset by real-time polymerase chain reaction (PCR) and antigen test. Of the tested SARS-CoV-2 positive sample pairs, 55.8% were identified as Omicron BA.1 and 44.2% as Omicron BA.2. Real-time PCR from buccal swabs generated significantly higher quantification cycle (Cq) values compared to those from matched combined oro-/nasopharyngeal swabs and resulted in an increased number of false-negative PCR results. Reduced diagnostic sensitivity of buccal swabs by real-time PCR was observed already at day one after symptom onset. Similarly, detection rates using the Abbott COVID-19 Ag Rapid Test Device were reduced in buccal swabs compared to those using combined oro-/nasopharyngeal swabs. Our results suggest reduced clinical diagnostic sensitivity of saliva from buccal swabs in comparison to combined oro-/nasopharyngeal swabs in the detection of Omicron in symptomatic individuals.

## Background

Respiratory tract specimens, such as those collected by oro-/nasopharyngeal swabs, are currently recommended for the direct detection of SARS-CoV-2 by real-time polymerase chain reaction (PCR) and most antigen tests [1–3]. In November 2021, a new SARS-CoV-2 variant B.1.1.529 (Omicron) emerged and spread rapidly around the globe [4]. Several studies have suggested an improved sensitivity of saliva over upper respiratory tract specimens in the detection of Omicron and other SARS-CoV-2 variants by real-time PCR [5–8]. Saliva could offer an appealing alternative to oro- and/or nasopharyngeal swabs as sample collection is considered less invasive and could potentially be easily performed by untrained caretakers and patients themselves [9]. Hence, the aim of this study was to assess the diagnostic performance of saliva versus oro-/nasopharyngeal swab samples in the detection of Omicron. To do so, we compared the clinical diagnostic sensitivity of matched buccal and combined oro-/nasopharyngeal swabs collected from hospitalized, symptomatic individuals in real-time PCR and antigen test.

## Methods

### Study design and sample collection

Clinical specimens were collected as part of the COViK study conducted by the Robert Koch Institute in collaboration with the Paul-Ehrlich-Institut. Thirteen hospitals across Germany served as study sites. Samples were collected between January and March 2022. Sampling was performed on symptomatic individuals on day six (median) after symptom onset. Trained study nurses performed sampling, using swabs of identical design (eSwab™, COPAN Diagnostics, Murrieta, CA, USA) for both buccal and combined oro-/nasopharyngeal sample collection. A total of 107 matched sample pairs consisting of one buccal and one combined oro-/nasopharyngeal swab were collected. Buccal swab collection was performed immediately before collection of the combined oro-/nasopharyngeal swab. Prior to buccal swabbing participants were asked to think of their favorite food for approximately 0.5-1.0 minute to stimulate saliva flow. Buccal swab samples were collected by streaking both the left and right lower inner cheek for 30 seconds each while applying light pressure to the swab and rotating it around its own axis allowing for full saturation of the swab tip with saliva. Subsequently, a fresh swab was used for oropharyngeal sampling directly followed by nasopharyngeal sampling using the same swab. After sample collection swabs were transferred to their respective collection tubes containing transport medium. Matched buccal and combined oro-/nasopharyngeal swabs were shipped together at 2-8°C and were stored at 4°C upon arrival, ensuring identical transport and storage conditions for matched samples. Time from sampling to result were 2 days (median), while nearly half of all samples (47.93 %) required only 1 day from sampling to result.

### RNA extraction and real-time PCR analysis

To extract viral RNA from samples, 140 µl of swab-containing transport medium were manually inactivated using AVL+Ethanol and extracted using the QIAamp Viral RNA Mini kit (QIAGEN, Hilden, Germany) according to manufacturer’s instructions. SARS-CoV-2 RNA was detected by real-time PCR using two separate assays, each targeting a distinct SARS-CoV-2 genomic region (E-gene and Orf1ab) as has been described previously [10]. For the identification of SARS-CoV-2 variants, PCR positive samples were further analyzed using variant-specific PCR assays and/or next-generation sequencing.

### Antigen testing

For antigen detection, the Panbio™ COVID-19 Ag Rapid Test Device (Abbott Rapid Diagnostics, Jena, Germany) was used. For testing, 50 µl of the native swab-containing transport medium were transferred directly to the test-specific extraction buffer and subsequent testing was performed according to manufacturer’s instructions. Results were analyzed independently by two trained laboratory technicians. If results were not in agreement a third person analyzed the test and the result in favor was noted. All antigen tests included in this study showed a visible control line.

## Results

First, we compared clinical diagnostic sensitivities for the detection of Omicron from matched buccal and combined oro-/nasopharyngeal swabs by real-time PCR. In total, 107 matched sample pairs were collected at median six days after symptom onset from previously confirmed SARS-CoV-2 positive hospitalized, symptomatic individuals. Of those, 11 sample pairs (10.28 %) tested PCR negative for SARS-CoV-2 in both buccal and oro-/nasopharyngeal swabs. Of the positive samples, all were identified as Omicron (55.8% BA.1 and 44.2% BA.2). Only two oro-/nasopharyngeal swabs (1.87 %) tested PCR negative while the matched buccal swabs tested PCR positive (Figure 1A). In contrast, 17 buccal swabs (15.89 %) tested PCR negative, while the matched oro-/nasopharyngeal swabs tested PCR positive, resulting in a higher number of false-negative real-time PCR results for buccal swabs in comparison to combined oro-/nasopharyngeal swabs (Figure 1A). Comparing only those sample pairs that tested PCR positive for both buccal and oro-/nasopharyngeal swab, real-time PCR from buccal swabs resulted in significantly higher Cq values compared to matching oro-/nasopharyngeal swabs with a difference in means for E-gene of 7.36 Cq (CI 6.23 to 8.5) and Orf1ab of 7.2 Cq (CI 6.1 to 8.3) (Figure 1B). Overall, lower Cq values in buccal swabs were observed for only 7 (E-gene) and 8 (Orf1ab) sample pairs. Notably, reduced performance of buccal swabs was observed for both Omicron BA.1 and BA.2 (data available upon request). Higher Cq values in buccal swabs were detected as early as day one to two after symptom onset (Figure 2). We also tested detection performance of matched buccal and combined oro-/nasopharyngeal swabs with the Abbott COVID-19 Ag Rapid Test Device. While the positive detection rate for combined oro-/nasopharyngeal swab samples by antigen test was 58.44 % (45/77 samples), positive detection rate for buccal swab samples was only 3.9 % (3/77 samples) (Figure 3).

**Figure 1.**
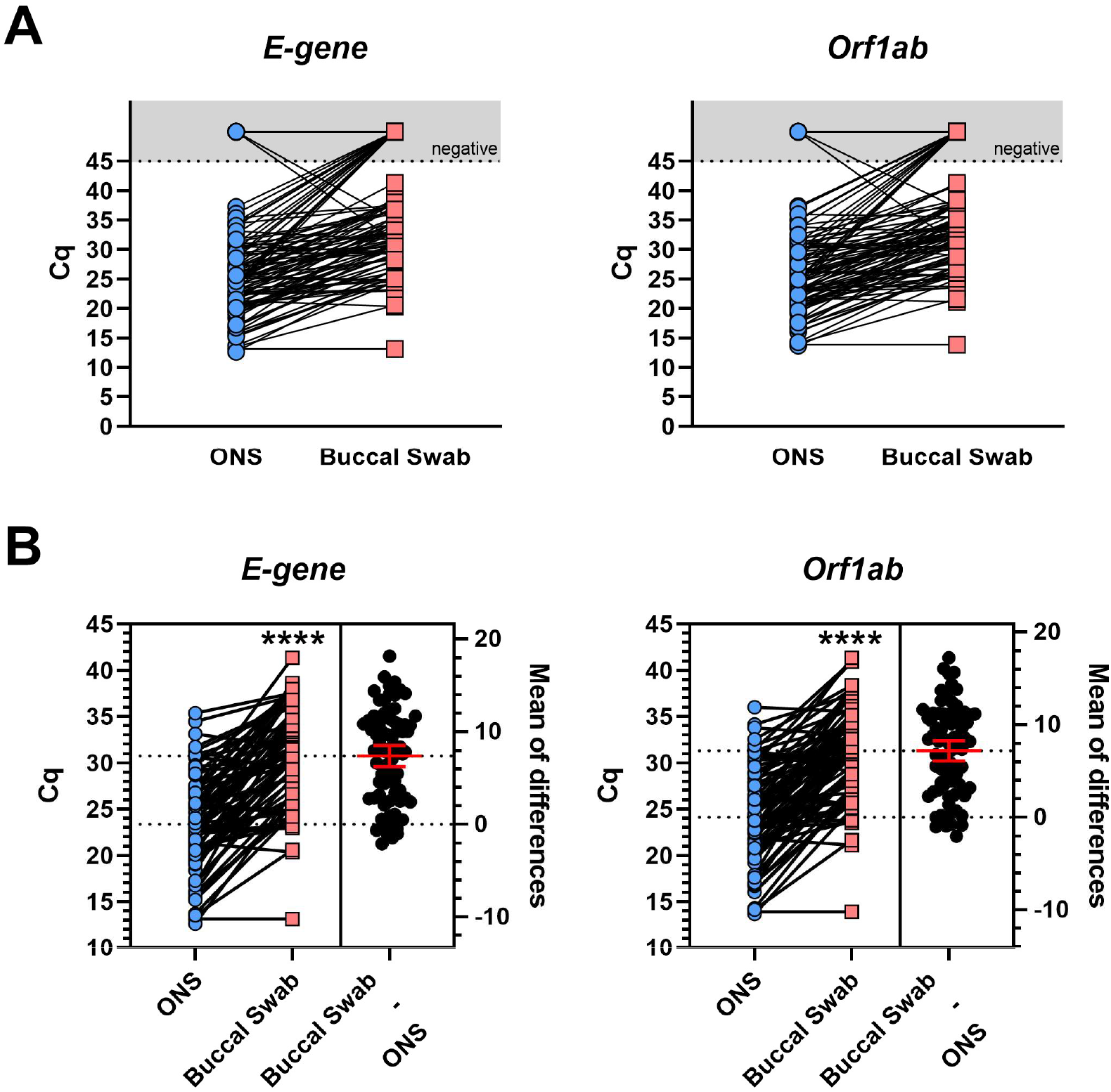
Comparison of SARS-CoV-2 B.1.1.529 Omicron detection performance of matched buccal and combined oro-/nasopharyngeal swabs by real-time PCR. **A** Comparison of cycle threshold (Cq) values for two distinct genomic regions of SARS-CoV-2 (E-gene and Orf1ab) of matched buccal and combined oro-/nasopharyngeal swabs (ONS) by quantitative real-time PCR (RKI/ZBS1 SARS-CoV-2 protocol, Michel *et al*. Virol J (2021) 18:110); n=107. **B** Estimation plot of SARS-CoV-2 positive sample pairs with Cq values ≤45; n=77. Line at mean with 95% Confidence Interval; Paired t test, p **** <0,0001.

**Figure 2.**
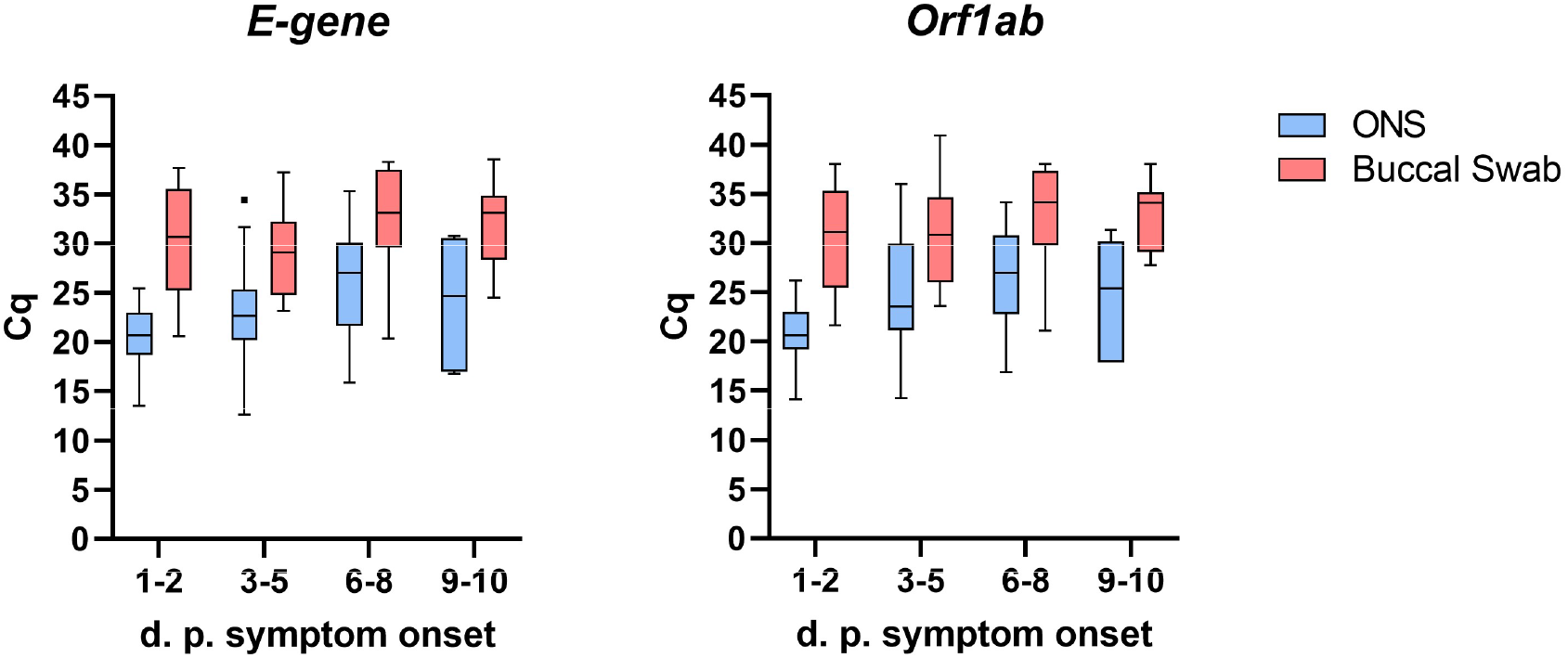
Comparison of real-time PCR results from buccal and combined oro-/nasopharyngeal swabs at different time periods after symptom onset. Comparison of cycle threshold (Cq) values for two distinct genomic regions of SARS-CoV-2 (E-gene and Orf1ab) of matched buccal and combined oro-/nasopharyngeal swabs (ONS) by quantitative real-time PCR (RKI/ZBS1 SARS-CoV-2 protocol, Michel *et al*. Virol J (2021) 18:110) at different days post (d.p.) symptom onset. Data displayed as Tukey box plot.

**Figure 3.**
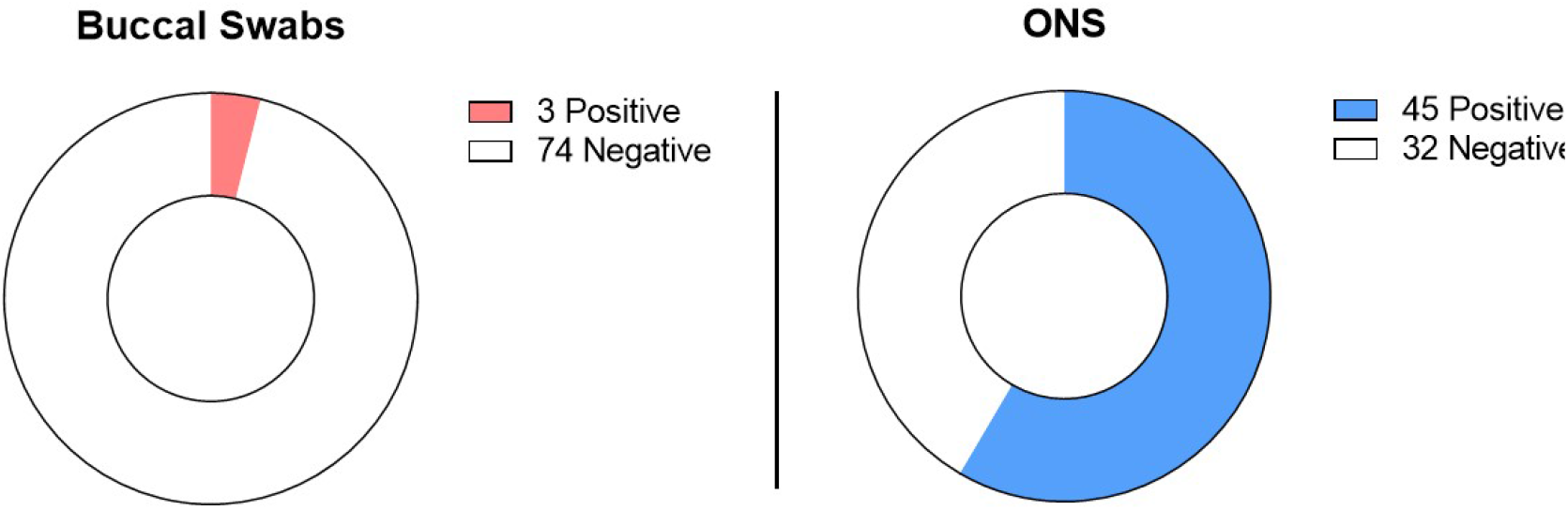
Comparison of SARS-CoV-2 B.1.1.529 Omicron detection performance of matched buccal and combined oro-/nasopharyngeal swabs by antigen test. PCR positive buccal and combined oro-/nasopharyngeal (ONS) sample pairs (n=77) were tested using the Panbio™ COVID-19 Ag Rapid Test Device. Shown are the number of positive and negative antigen test results. All tests showed a visible control line.

## Discussion

In this study, we observed reduced clinical diagnostic sensitivity of saliva collected by buccal swabs in comparison to matched combined oro-/nasopharyngeal swabs in the detection of Omicron (BA.1 and BA.2). Serval studies on the sensitivity of saliva versus respiratory tract specimens for the detection of SARS-CoV-2, including Omicron, have been conducted, leading to mixed and in parts contradictory results [5–8,11–13]. In this study, samples were collected from hospitalized, symptomatic individuals who had previously been confirmed to be SARS-CoV-2 positive, resulting in sample collection at median six days after initial symptom onset. We observed that around 10 % of initially PCR positive individuals were negative by the time of the second PCR testing, probably due to the relatively late time of sampling. Despite the majority of samples being collected at late stages of infection, higher Cq values in buccal swabs were detected already from day one to two after symptom onset in this limited data set. In a recent study, Lai *et al*. compared saliva and nasal swabs from close contacts of COVID-19 cases over time and found that, in those contacts who became infected, saliva samples showed higher viral loads compared to those in nasal swabs from three days prior to symptom onset to two days after symptom onset [14]. In contrast, two days after symptom onset there was a trend towards improved sensitivity with nasal swabs compared to saliva, indicating the importance of time of sampling for subsequent specimen sensitivity [14]. Furthermore, we applied buccal swabbing to collect saliva using swabs of identical design for both the collection of saliva and oro-/nasopharyngeal specimens. Using identical swabs enabled direct comparison between specimen types by ensuring identical conditions for transport and handling during all downstream manipulations, including RNA extraction. However, we did not assess other types of swabs and saliva sampling methods, such as drooling, spitting or sampling from specific salivary glands or other locations, which might impact subsequent saliva sensitivity. Overall, factors such as the time of sampling and specific sampling methods are likely to play a critical role in the diagnostic sensitivity of saliva and might explain some of the differences found across studies.

In addition to real-time PCR, we also used the Abbott Panbio™ COVID-19 Ag Rapid Test for the detection of Omicron by viral antigen, which resulted in substantially reduced detection rates among buccal swabs in comparison to combined oro-/nasopharyngeal swabs. While reduced performance of this antigen test for the detection of Omicron is not predicted by the manufacturer due to the use of the nucleocapsid (N) protein as target antigen [15], it has previously been shown that the use of throat and saliva samples with the Panbio™ COVID-19 Ag Rapid Test led to poorer sensitivity compared to nasopharyngeal swab samples [16]. Although all swab samples in this study were subject to dilution in transport medium as well as the antigen test extraction buffer, it is not clear whether the reduced performance of buccal swab samples is due to a reduced concentration of N protein in buccal saliva or whether saliva is a suboptimal sample type for use in the Abbott Panbio™ COVID-19 Ag Rapid Test.

At the time of study, Omicron BA.1 and BA.2 were the dominant variants present in Germany [17], which is also reflected in our sample set. A study using *ex vivo* infections of different tissues found that Omicron BA.2 displayed increased replication competence in human nasal and bronchial tissues compared to Omicron BA.1 as well as the original SARS-CoV-2 wild-type strain and the Delta variant [18]. It remains to be elucidated how recently emerged and currently dominant Omicron variants affect diagnostic sensitivities of different specimen types.

Taken together, despite the reduced invasiveness and ease of sampling, the use of saliva collected by buccal swabs displays substantially reduced sensitivity in comparison to combined oro-/nasopharyngeal swab specimens for the detection of Omicron. This further highlights the importance to carefully consider time and context of sampling for choosing the optimal specimen type for diagnostics.

## Data Availability

All data produced in the present work are either contained in the manuscript or are available upon reasonable request to the authors.

## Conflict of interest

Authors declared no conflict of interest.

## Ethical statement

The study obtained ethical approval by the Berliner Ärztekammer (Berlin Chamber of Physicians, Eth 20/40).

## Acknowledgements

The authors thank all study nurses for the valuable contribution, namely Sawsanh Al-Ogaidi, Nancy Beetz, Belgin Esen, Rola Khalife, Katja Lange, Luise Mauer, Antje Micheel, Marlies Schmidt, Yvonne Weis, Franziska Weiser and Aysete Yencilek. The authors are grateful to Ursula Erikli for copy-editing. This work was funded by the German Federal Ministry of Health.

## Notes

### Competing Interest Statement

The authors have declared no competing interest.

### Author Declarations

The study obtained ethical approval by the Berliner Aerztekammer (Berlin Chamber of Physicians Eth, 20/40).

